# Fine mapping of HLA effects in Oral and non-Oral lichen planus

**DOI:** 10.1101/2025.11.17.25340396

**Authors:** Jarmo Ritari, Mary Reeve, FinnGen, Maria Siponen, Mari Vehviläinen, Tuula Salo, Kazutoyo Osoegawa, Marcelo Fernandez Viña, Benjamin Goudey, Jukka Partanen, Emmanuel JM Mignot

**Author notes:** <u>Correspondence should be addressed to:</u> Prof. Emmanuel Mignot, Stanford School of Medicine, 3165 Porter Drive, #2178, Palo Alto CA 94304, USA, Or Prof. Jukka Partanen, Research and Development, Finnish Red Cross Blood, Service, Helsinki, Finland. Participants of the FinnGen are listed in Supplementary Table 1.

## Abstract

Lichen planus (LP) is an inflammatory disease affecting squamous epithelia, typically manifesting in a cutaneous (non-OLP) and an oral mucosa (OLP) form, the latter conferring elevated risk of oral squamous cell carcinoma. Despite presence of CD4+ and CD8+ T-cell infiltrates in LP lesions, specific autoantibodies or target T-cell antigens have not been identified. A recent genome wide association study (GWAS) uncovered 27 genome-wide independent associations, with the strongest signal within HLA class II, particularly involving *DQB1*05:01*. This association showed stronger effects in non-OLP (OR=2.09) versus OLP (OR=1.36).

Here, we performed a high-resolution HLA fine-mapping analysis in FinnGen to dissect this strong class II signal and clarify its relationship to LP subtypes. We find that most *DQA1*01∼DQB1*05:01* haplotypes confer increased susceptibility, with the highest risk conferred by *DQA1*01:05∼DQB1*05:01* followed by *DQA1*01:01∼DQB1*05:01*. In subjects not carrying *DQB1*05:01*, *DRB1*15:01∼DQA1*01:02∼DQB1*06:02* had a strong protective effect, more pronounced in non-OLP than OLP. Further associations were found in *DRB1*09:01* and *DQB1*02:02* as well as independent HLA class I associations with *A*03:01*, *B*08:01* and *B*13:02*, all stronger in OLP versus non-OLP. Conditioning SNP associations for these effects eliminated the HLA GWAS signal.

These effects highlight that while the *DQB1*05:01* association remains largely invariant to *DQA1* polymorphisms across LP subtypes, the variable strength of HLA associations in non-OLP and OLP forms suggests distinct immunopathological mechanisms. The presence of trans-heterodimer effects in this disease illustrates the need to analyze *HLA-DQ*-associated diseases with methods beyond simple conditioning. The invariance to *DQA1* polymorphisms may facilitate the identification of potential pathological epitopes.

## Introduction

Lichen planus (LP) is a chronic inflammatory mucocutaneus disease of unknown origin that causes lesions and irritation on the skin (cutaneous LP), mucous membranes (penile, vulvar, oral, or esophageal LP), scalp (lichen planopilaris), or nails (lichen unguis), with a lifelong prevalence of ∼1% of the population(1, 2). Most common forms of LP are oral (OLP) and cutaneous LP (non-OLP). OLP is considered more significant, as it tends to be chronic and is associated with an increased risk of squamous cell carcinoma(3). Non-OLP frequently leaves hyperpigmentation marks upon resolution and may relapse. As LP lesions demonstrate CD4+ and CD8+ T cell infiltrates(4), and no infectious agent has been systematically found (although an association with hepatitis C virus (HCV) has been documented(5–7), it is considered a T-cell mediated autoinflammatory disease.

Until recently, limited genetic studies had been conducted in LP. This changed with the study of Reeve *et al*.(8) who conducted genome-wide association analyses (GWAS) in FinnGen on OLP (n = 3,323) and non-OLP (n = 4,356) cases with replication in the UK Biobank (UKBB), where subtype information was unavailable and thus combined for replication. These authors identified 27 independent associations at 25 distinct genomic locations, many of which were shared between OLP and non-OLP, with seven associations unique to one subgroup. Most of these associations overlapped with other autoimmune diseases, most notably hypothyroidism. Beyond the autoimmune disease overlap, a notable finding was the association of nine SNPs with increased eosinophil counts, potentially indicating a previously underappreciated role of these cells in LP pathogenesis(8). The most significant association noted was in the HLA complex, with imputation suggesting a primary association with *DQB1*05:01* (p=7.5×10^−211^), an effect that was stronger in non-OLP vs OLP (OR=2.09 vs 1.36)(8).

The analysis of HLA is complicated by the potential role of DQ trans-heterozygote effects, which have been identified in other strongly HLA-DQ associated diseases such as celiac disease and narcolepsy (9–12). In some cases, novel heterodimers formed between α and β chains encoded on different chromosomes may increase disease susceptibility(12). In other cases, trans-encoded heterodimers may be functionally inactive and reduce susceptibility by competing for heterodimerization with the primary susceptibility heterodimer encoded in cis(10, 11). Indeed, trans DQ heterodimers can also influence disease predisposition or in transplantation outcomes including rejection, GvHD or disease relapse(13, 14), although these trans-complementations can only occur within the DQ1 (*DQA1*01* bearing haplotypes) and within non-DQ1 (*DQA1*02*, *03* or *04* bearing haplotypes) families because of molecular constrains for subunit pairing.

In our current study, we performed a fine-mapping HLA analysis of the strong class II signal detected in the previous GWAS to clarify the varying strength patterns of allele association in LP subtypes. We specifically analyzed possible trans-heterodimer effects in the *DQ* locus. Our results demonstrate that the previous *DQB1*05:01* association represents the collective influence of multiple *HLA-DQ* and Class I effects, and that *DQB1*06:02* confers and independent protective effect. With a few exceptions, we confirmed that the HLA signal is highly similar between non-OLP and OLP, differing primarily in magnitude, suggesting etiological overlap in non-OLP vs OLP, as suggested by the GWAS study. Interestingly, although most HLA associations were stronger in non-OLP, several associations were unique to OLP, suggesting distinct HLA factors may be involved in OLP pathogenesis.

## Methods

### Study cohort

The FinnGen study(15) (www.finngen.fi), is a large-scale genomics initiative that has analyzed over 500,000 Finnish biobank samples and correlated genetic variation with health data to understand disease mechanisms and predispositions. The project is a collaboration between research organisations and biobanks within Finland and international industry partners (Supplementary Table 1). The FinnGen includes biobank participants with acute and chronic diseases, healthy blood donors, and population collections. FinnGen data freeze 12 (DF12) consists of 520,210 individuals. We used the deep electronic health registry data in FinnGen DF12 and records of clinic of diagnosis(8) to create OLP(n=3,651) and non-OLP (n=4,808) subgroupings. 490,211 individuals without an LP ICD diagnosis were used as controls.

### Phenotypic definitions

Individuals with LP were identified using ICD codes: ICD10 L43 (with extensions 0, 1, 3, 9) and ICD8/9 6970 (with Finnish extensions A, B, C). Due to the lack of body location specificity in these codes, patients were categorized into subgroups of oral LP (OLP) and non-oral LP (non-OLP) based on the clinic type where the diagnosis was made. Specifically, diagnoses made at dental or orthodontic clinics were classified as OLP, while all others were classified as non-OLP, following the methodology of Reeve and co-workers(8).

### HLA data and association analysis

Array-based genotype data in FinnGen were called and subjected to variant and sample level quality control (QC) followed by phasing and imputation as described in the FinnGen flagship paper(15). HLA haplotypes are denoted by alleles separated by “∼”. HLA heterodimers are denoted by alleles separated by “/”.

Alleles of classical HLA genes (*A, B, C, DRB1, DQA1, DQB1, and DPB1*) were imputed as part of the FinnGen analysis pipeline using a Finnish population-specific reference panel(16) and the HIBAG algorithm(17). Posterior probability (PP) of imputation was retained in the HLA allele dosage information within the GP field of VCF data that were produced by the imputation pipeline (https://github.com/FRCBS/HLA-imputation/tree/master/src) using plink2 command --vcf dosage=GP.

We performed conditional association analyses by selecting a specific HLA allele dosage as an additional analysis covariate. To select specific combinations of DQ alleles for analysis of DQ haplotypes, we handled the imputed HLA alleles as integer dosage values in R. Thresholds for converting imputed dosages to integer values were as follows: <0.5 (0), ≥0.5 & <1.5 (1), and ≥1.5 (2). We then converted the allele data to plink genotype format (.bed) by first transforming the dosages into ped/map text files using R. We applied the plink command --make-bed for this conversion. *DQA1∼DQB1* haplotype homozygotes were defined as individuals with an imputed allele dosage of ≥1.5 in both genes simultaneously. We coded the resulting allele combinations as a binary variable, depending on the homozygosity status of the alleles. These genotypes were then converted to PLINK format as described above.

### Rationale for analyzing complex HLA effects

To properly dissect the complex effects of multiple HLA alleles in lichen planus, multiple parallel analyses were necessary. Initially, effects in class I and class II regions were analyzed independently and conditioned on each other to identify linkage disequilibrium (LD) effects. Relative predisposition effects, i.e., displacement of alleles other than the main risk allele, are identified by conditioning or sequentially removing selected genotypes(18) and analyzing residual effects, especially in cases of trans-heterodimer effects in DQ-associated diseases. Simple conditioning may be inadequate, requiring the removal of specific genotypes. We employed also a homozygous only analysis where all other samples than DQ homozygotes are excluded. This allows observing the main DQ effects without the complication of trans-heterodimer effects. These complex effects necessitate understanding allele homology and amino acid polymorphisms, particularly those in exon 3 (encoding the HLA α2/β2 domain) or the signal peptide (Supplementary Figure 1), which may not affect peptide binding and presentation, unlike those in exon 1 and 2. The analyses involve iterative processes and evaluation of the results in the light of the DQ effects described above. Supplementary Tables 2 and 3 and Supplementary Figure 2 detail the designs and analyses used, while Supplementary Figure 3 reports on LD between associated alleles. Supplementary Table 4 shows what are likely possible DQα, DQβ allele pairings based on their presence in known, well documented haplotypes.

### Data analysis

Data management and visualization was conducted using R software version 4.3.2 or later (https://www.r-project.org/) with the packages tidyverse(19) version 1.3.0 and data.table(20) version 1.15.4 in RStudio. For association analyses, we used Regenie(21) version 3.0.1 or 3.6 with default settings, taking plink-formatted HLA genotypes as input. Genotype data for the full genome null model in Regenie step 1 were prepared using FinnGen HapMap3 variants in plink format. To select a set of independent common SNPs for the null model, we applied additional filters using plink(22) commands: --maf 0.1, --mac 100, --geno 0.1, --hwe 1e-15, --mind 0.01, and --thin-count 500000.

Our analysis included the basic covariates: age at death or end of follow-up, sex, sampling year, region of birth, FinnGen2 chip, batch, and the first ten genetic principal components. Conditional analyses incorporated HLA allele dosages as additional covariates, depending on the conditioning step.

To account for multiple testing, we corrected association p-values across tested HLA genotypes and the three LP phenotypes (OLP, non-OLP, All) using the Benjamini-Yekutieli method(23). A false discovery rate (FDR) threshold of <0.05 was considered significant. Detailed analysis results presented in the figures are reported as supplementary data.

### Ethics Statement

Study subjects in FinnGen provided informed consent for biobank research, based on the Finnish Biobank Act. Alternatively, separate research cohorts, collected prior the Finnish Biobank Act came into effect (in September 2013) and start of FinnGen (August 2017), were collected based on study-specific consents and later transferred to the Finnish biobanks after approval by Fimea (Finnish Medicines Agency), the National Supervisory Authority for Welfare and Health. Recruitment protocols followed the biobank protocols approved by Fimea. The Coordinating Ethics Committee of the Hospital District of Helsinki and Uusimaa (HUS) statement number for the FinnGen study is Nr HUS/990/2017.

The FinnGen study is approved by Finnish Institute for Health and Welfare (permit numbers: THL/2031/6.02.00/2017, THL/1101/5.05.00/2017, THL/341/6.02.00/2018, THL/2222/6.02.00/2018, THL/283/6.02.00/2019, THL/1721/5.05.00/2019 and THL/1524/5.05.00/2020), Digital and population data service agency (permit numbers: VRK43431/2017-3, VRK/6909/2018-3, VRK/4415/2019-3), the Social Insurance Institution (permit numbers: KELA 58/522/2017, KELA 131/522/2018, KELA 70/522/2019, KELA 98/522/2019, KELA 134/522/2019, KELA 138/522/2019, KELA 2/522/2020, KELA 16/522/2020), Findata permit numbers THL/2364/14.02/2020, THL/4055/14.06.00/2020, THL/3433/14.06.00/2020, THL/4432/14.06/2020, THL/5189/14.06/2020, THL/5894/14.06.00/2020, THL/6619/14.06.00/2020, THL/209/14.06.00/2021, THL/688/14.06.00/2021, THL/1284/14.06.00/2021, THL/1965/14.06.00/2021, THL/5546/14.02.00/2020, THL/2658/14.06.00/2021, THL/4235/14.06.00/2021, Statistics Finland (permit numbers: TK-53-1041-17 and TK/143/07.03.00/2020 (earlier TK-53-90-20) TK/1735/07.03.00/2021, TK/3112/07.03.00/2021) and Finnish Registry for Kidney Diseases permission/extract from the meeting minutes on 4^th^ July 2019.

The Biobank Access Decisions for FinnGen samples and data utilized in FinnGen Data Freeze 11 include: THL Biobank BB2017_55, BB2017_111, BB2018_19, BB_2018_34, BB_2018_67, BB2018_71, BB2019_7, BB2019_8, BB2019_26, BB2020_1, BB2021_65, Finnish Red Cross Blood Service Biobank 7.12.2017, Helsinki Biobank HUS/359/2017, HUS/248/2020, HUS/430/2021 §28, §29, HUS/150/2022 §12, §13, §14, §15, §16, §17, §18, §23, §58, §59, HUS/128/2023 §18, Auria Biobank AB17-5154 and amendment #1 (August 17 2020) and amendments BB_2021-0140, BB_2021-0156 (August 26 2021, Feb 2 2022), BB_2021-0169, BB_2021-0179, BB_2021-0161, AB20-5926 and amendment #1 (April 23 2020) and it’s modifications (Sep 22 2021), BB_2022-0262, BB_2022-0256, Biobank Borealis of Northern Finland_2017_1013, 2021_5010, 2021_5010 Amendment, 2021_5018, 2021_5018 Amendment, 2021_5015, 2021_5015 Amendment, 2021_5015 Amendment_2, 2021_5023, 2021_5023 Amendment, 2021_5023 Amendment_2, 2021_5017, 2021_5017 Amendment, 2022_6001, 2022_6001 Amendment, 2022_6006 Amendment, 2022_6006 Amendment, 2022_6006 Amendment_2, BB22-0067, 2022_0262, 2022_0262 Amendment, Biobank of Eastern Finland 1186/2018 and amendment 22§/2020, 53§/2021, 13§/2022, 14§/2022, 15§/2022, 27§/2022, 28§/2022, 29§/2022, 33§/2022, 35§/2022, 36§/2022, 37§/2022, 39§/2022, 7§/2023, 32§/2023, 33§/2023, 34§/2023, 35§/2023, 36§/2023, 37§/2023, 38§/2023, 39§/2023, 40§/2023, 41§/2023, Finnish Clinical Biobank Tampere MH0004 and amendments (21.02.2020 & 06.10.2020), BB2021-0140 8§/2021, 9§/2021, §9/2022, §10/2022, §12/2022, 13§/2022, §20/2022, §21/2022, §22/2022, §23/2022, 28§/2022, 29§/2022, 30§/2022, 31§/2022, 32§/2022, 38§/2022, 40§/2022, 42§/2022, 1§/2023, Central Finland Biobank 1-2017, BB_2021-0161, BB_2021-0169, BB_2021-0179, BB_2021-0170, BB_2022-0256, BB_2022-0262, BB22-0067, Decision allowing to continue data processing until 31^st^ Aug 2024 for projects: BB_2021-0179, BB22-0067,BB_2022-0262, BB_2021-0170, BB_2021-0164, BB_2021-0161, and BB_2021-0169, and Terveystalo Biobank STB 2018001 and amendment 25^th^ Aug 2020, Finnish Hematological Registry and Clinical Biobank decision 18^th^ June 2021, Arctic biobank P0844: ARC_2021_1001.

## Results

### Non-adjusted HLA association analysis reconfirms *DQB1*05:01* as key driver of LP susceptibility

A recent GWAS found a highly significant association peak in the HLA class II region extending into class I(8). To understand which HLA allele was primarily associated with lichen planus, we analyzed the association of all HLA alleles within *HLA-DR, DQ, DP, A, B* and *C* with LP overall (non-OLP+OLP), non-OLP only and OLP only (Table 1 and 2)

**Table 1.** HLA allele associations without additional covariates.

| HLA | Frequency | OLP |  |  | All |  |  | non-OLP |  |  |
| --- | --- | --- | --- | --- | --- | --- | --- | --- | --- | --- |
|  |  | P | OR | 95% CI | P | OR | 95% CI | P | OR | 95% CI |
| DQB1*05:01 | 0.194 | 1.64E-25 | 1.341 | 1.27 - 1.41 | 4.99E-208 | 1.714 | 1.66 - 1.77 | 1.71E-215 | 1.99 | 1.91 - 2.07 |
| DQA1*01:01 | 0.185 | 1.37E-24 | 1.342 | 1.27 - 1.42 | 9.57E-181 | 1.676 | 1.62 - 1.73 | 1.81E-182 | 1.922 | 1.85 - 2 |
| *DRB1\*01:01* | 0.179 | 2.10E-24 | 1.348 | 1.27 - 1.42 | 3.08E-177 | 1.682 | 1.63 - 1.74 | 3.46E-178 | 1.929 | 1.85 - 2.01 |
| *B\*35:01* | 0.122 | 1.42E-10 | 1.261 | 1.18 - 1.35 | 7.82E-77 | 1.527 | 1.46 - 1.59 | 1.51E-79 | 1.726 | 1.64 - 1.82 |
| *C\*04:01* | 0.146 | 5.89E-08 | 1.201 | 1.13 - 1.28 | 1.67E-65 | 1.443 | 1.39 - 1.5 | 2.24E-70 | 1.623 | 1.54 - 1.71 |
| *A\*03:01* | 0.224 | 1.19E-10 | 1.21 | 1.14 - 1.28 | 4.55E-49 | 1.327 | 1.28 - 1.38 | 2.21E-45 | 1.423 | 1.36 - 1.49 |
| *DQA1\*01:02* | 0.182 | 1.96E-20 | 0.737 | 0.69 - 0.79 | 3.68E-42 | 0.743 | 0.71 - 0.78 | 8.69E-24 | 0.75 | 0.71 - 0.79 |
| *DQB1\*06:02* | 0.139 | 1.93E-20 | 0.709 | 0.66 - 0.76 | 3.15E-39 | 0.725 | 0.69 - 0.76 | 7.54E-21 | 0.74 | 0.69 - 0.79 |
| *DRB1\*15:01* | 0.138 | 3.06E-20 | 0.711 | 0.66 - 0.77 | 3.88E-38 | 0.729 | 0.69 - 0.77 | 6.15E-20 | 0.746 | 0.7 - 0.8 |
| *DPB1\*04:02* | 0.188 | 2.81E-07 | 1.168 | 1.1 - 1.24 | 4.86E-35 | 1.274 | 1.23 - 1.32 | 3.25E-32 | 1.35 | 1.29 - 1.42 |
| *DQA1\*01:05* | 0.008 | 6.24E-02 | 1.303 | 0.99 - 1.72 | 5.92E-31 | 2.305 | 2.03 - 2.62 | 4.46E-39 | 3.06 | 2.65 - 3.53 |
| *DRB1\*10:01* | 0.008 | 7.06E-02 | 1.282 | 0.98 - 1.68 | 1.13E-30 | 2.24 | 1.98 - 2.53 | 5.35E-39 | 2.96 | 2.57 - 3.41 |
| *C\*07:02* | 0.139 | 2.95E-04 | 0.88 | 0.82 - 0.94 | 3.29E-21 | 0.794 | 0.76 - 0.83 | 8.61E-21 | 0.737 | 0.69 - 0.79 |
| *DRB1\*13:01* | 0.095 | 4.51E-08 | 0.788 | 0.72 - 0.86 | 1.43E-18 | 0.774 | 0.73 - 0.82 | 6.30E-12 | 0.768 | 0.71 - 0.83 |
| *DQB1\*03:01* | 0.108 | 1.90E-04 | 0.863 | 0.8 - 0.93 | 9.28E-17 | 0.796 | 0.75 - 0.84 | 9.15E-16 | 0.745 | 0.69 - 0.8 |
| *DQA1\*01:03* | 0.09 | 7.43E-07 | 0.801 | 0.73 - 0.88 | 2.37E-16 | 0.782 | 0.74 - 0.83 | 4.30E-11 | 0.77 | 0.71 - 0.83 |
| *DQB1\*06:03* | 0.09 | 6.96E-07 | 0.801 | 0.73 - 0.88 | 3.72E-16 | 0.784 | 0.74 - 0.83 | 7.42E-11 | 0.773 | 0.71 - 0.84 |
| *DQA1\*05:05* | 0.068 | 1.38E-05 | 0.796 | 0.72 - 0.88 | 3.88E-16 | 0.75 | 0.7 - 0.81 | 2.96E-13 | 0.709 | 0.64 - 0.78 |
| *DPB1\*05:01* | 0.023 | 1.20E-04 | 0.734 | 0.63 - 0.86 | 4.93E-15 | 0.624 | 0.55 - 0.71 | 2.67E-11 | 0.583 | 0.49 - 0.69 |
| *DRB1\*11:01* | 0.035 | 1.76E-04 | 0.767 | 0.67 - 0.88 | 2.25E-14 | 0.672 | 0.6 - 0.75 | 1.59E-12 | 0.61 | 0.53 - 0.71 |
| *DRB1\*08:01* | 0.095 | 3.00E-06 | 0.819 | 0.75 - 0.89 | 2.12E-13 | 0.812 | 0.77 - 0.86 | 6.40E-09 | 0.805 | 0.75 - 0.87 |
| *DQA1\*04:01* | 0.097 | 2.83E-06 | 0.822 | 0.76 - 0.89 | 3.25E-13 | 0.817 | 0.77 - 0.86 | 1.01E-08 | 0.811 | 0.75 - 0.87 |
| *DQB1\*04:02* | 0.096 | 3.22E-06 | 0.822 | 0.76 - 0.89 | 3.50E-13 | 0.816 | 0.77 - 0.86 | 9.89E-09 | 0.81 | 0.75 - 0.87 |
| *B\*07:02* | 0.121 | 9.04E-03 | 0.903 | 0.84 - 0.97 | 1.11E-11 | 0.834 | 0.79 - 0.88 | 2.38E-11 | 0.789 | 0.73 - 0.85 |
| *C\*05:01* | 0.05 | 3.02E-05 | 0.772 | 0.68 - 0.88 | 7.03E-10 | 0.776 | 0.71 - 0.84 | 1.19E-05 | 0.789 | 0.71 - 0.88 |
| *DRB1\*04:01* | 0.09 | 3.17E-01 | 0.956 | 0.87 - 1.04 | 1.45E-09 | 0.829 | 0.78 - 0.88 | 1.53E-12 | 0.743 | 0.68 - 0.81 |
| *DQB1\*03:02* | 0.118 | 4.79E-01 | 0.974 | 0.9 - 1.05 | 4.49E-09 | 0.861 | 0.82 - 0.91 | 2.46E-12 | 0.786 | 0.73 - 0.84 |
| *A\*68:01* | 0.058 | 2.45E-04 | 0.824 | 0.74 - 0.91 | 7.57E-09 | 0.808 | 0.75 - 0.87 | 8.43E-06 | 0.805 | 0.73 - 0.89 |
| *B\*44:02* | 0.056 | 4.84E-06 | 0.758 | 0.67 - 0.86 | 9.29E-09 | 0.796 | 0.74 - 0.86 | 4.18E-04 | 0.839 | 0.76 - 0.92 |
| *DQA1\*03:01* | 0.11 | 7.06E-01 | 0.985 | 0.91 - 1.07 | 1.06E-08 | 0.854 | 0.81 - 0.9 | 1.13E-12 | 0.767 | 0.71 - 0.83 |

**Table 2.**
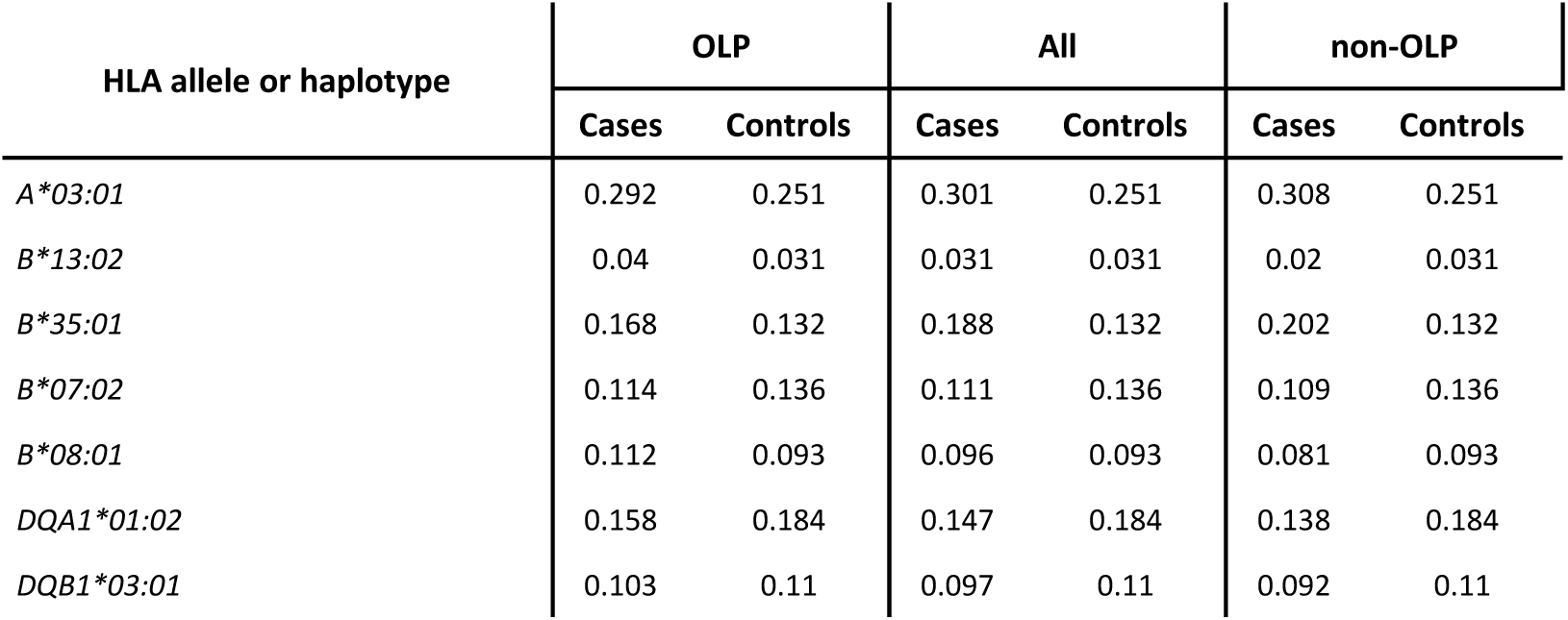
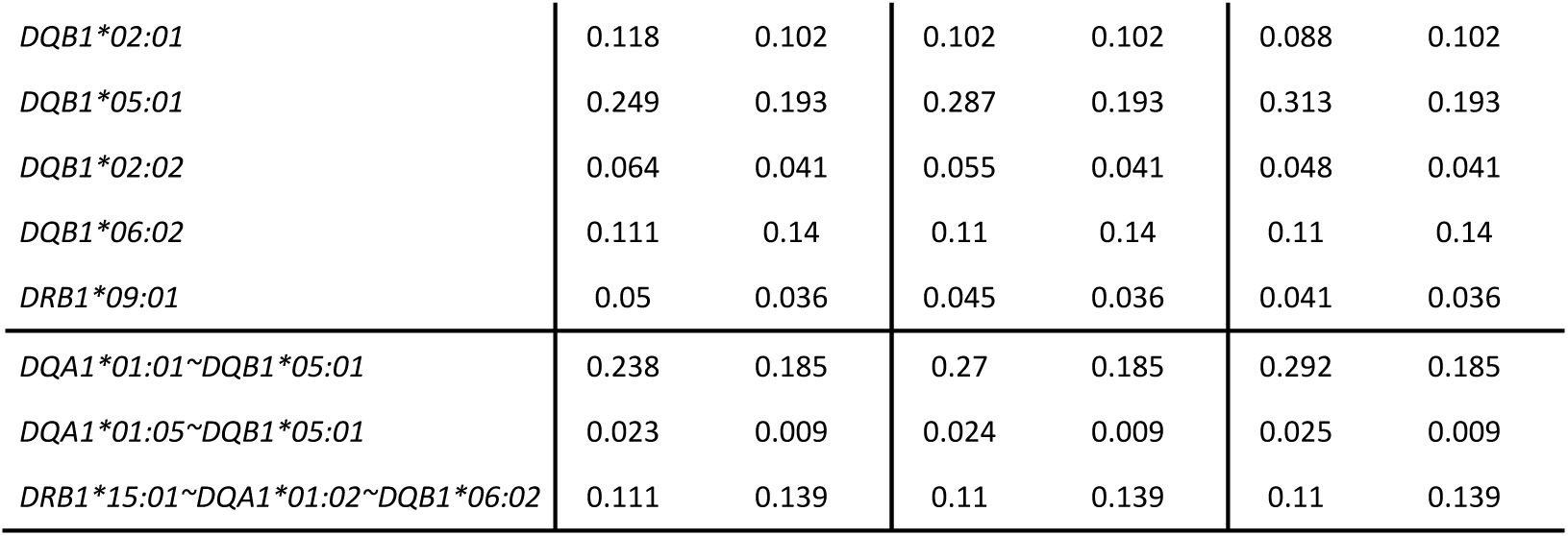
Case–control comparison of HLA allele frequencies. For haplotypes, the reported frequency represents an estimate derived from all constituent alleles and may differ from the frequencies of individual alleles.

| HLA allele or haplotype | OLP |  | All |  | non-OLP |  |
| --- | --- | --- | --- | --- | --- | --- |
|  | Cases | Controls | Cases | Controls | Cases | Controls |
| *A\*03:01* | 0.292 | 0.251 | 0.301 | 0.251 | 0.308 | 0.251 |
| *B\*13:02* | 0.04 | 0.031 | 0.031 | 0.031 | 0.02 | 0.031 |
| *B\*35:01* | 0.168 | 0.132 | 0.188 | 0.132 | 0.202 | 0.132 |
| *B\*07:02* | 0.114 | 0.136 | 0.111 | 0.136 | 0.109 | 0.136 |
| *B\*08:01* | 0.112 | 0.093 | 0.096 | 0.093 | 0.081 | 0.093 |
| *DQA1\*01:02* | 0.158 | 0.184 | 0.147 | 0.184 | 0.138 | 0.184 |
| *DQB1\*03:01* | 0.103 | 0.11 | 0.097 | 0.11 | 0.092 | 0.11 |
| *DQB1\*02:01* | 0.118 | 0.102 | 0.102 | 0.102 | 0.088 | 0.102 |
| *DQB1\*05:01* | 0.249 | 0.193 | 0.287 | 0.193 | 0.313 | 0.193 |
| *DQB1\*02:02* | 0.064 | 0.041 | 0.055 | 0.041 | 0.048 | 0.041 |
| *DQB1\*06:02* | 0.111 | 0.14 | 0.11 | 0.14 | 0.11 | 0.14 |
| *DRB1\*09:01* | 0.05 | 0.036 | 0.045 | 0.036 | 0.041 | 0.036 |
| *DQA1\*01:01~DQB1\*05:01* | 0.238 | 0.185 | 0.27 | 0.185 | 0.292 | 0.185 |
| *DQA1\*01:05~DQB1\*05:01* | 0.023 | 0.009 | 0.024 | 0.009 | 0.025 | 0.009 |
| *DRB1\*15:01~DQA1\*01:02~DQB1\*06:02* | 0.111 | 0.139 | 0.11 | 0.139 | 0.11 | 0.139 |

Consistent with Reeve *et al*.(8), the strongest association overall was with *DQB1*05:01* and alleles *DQA1*01:01* and *DRB1*01:01*, known to form the most common HLA class II haplotype in Finland. HLA class I *A*03:01*, *B*35:01*, *C*04:01* were also significant (Table 1). These above-mentioned HLA class I and II alleles form the most common HLA haplotype in Finland. Following this, significant *DRB1*15:01∼ DQA1*01:02∼DQB1*06:02* associations highlight a likely independent, protective signal(24) (Figure 1). *DRB1*10:01∼DQA1*01:05∼DQB1*05:01* was also significantly associated with a high odds ratio (Table 1 and 2). This is expected given it is similar in sequence to *DRB1*01:01∼DQA1*01:01∼DQB1*05:01* in the DQ region but rarer (*DQA1*01:01* and *DQA1*01:05* only differ by two amino acids, see below).

**Figure 1.**
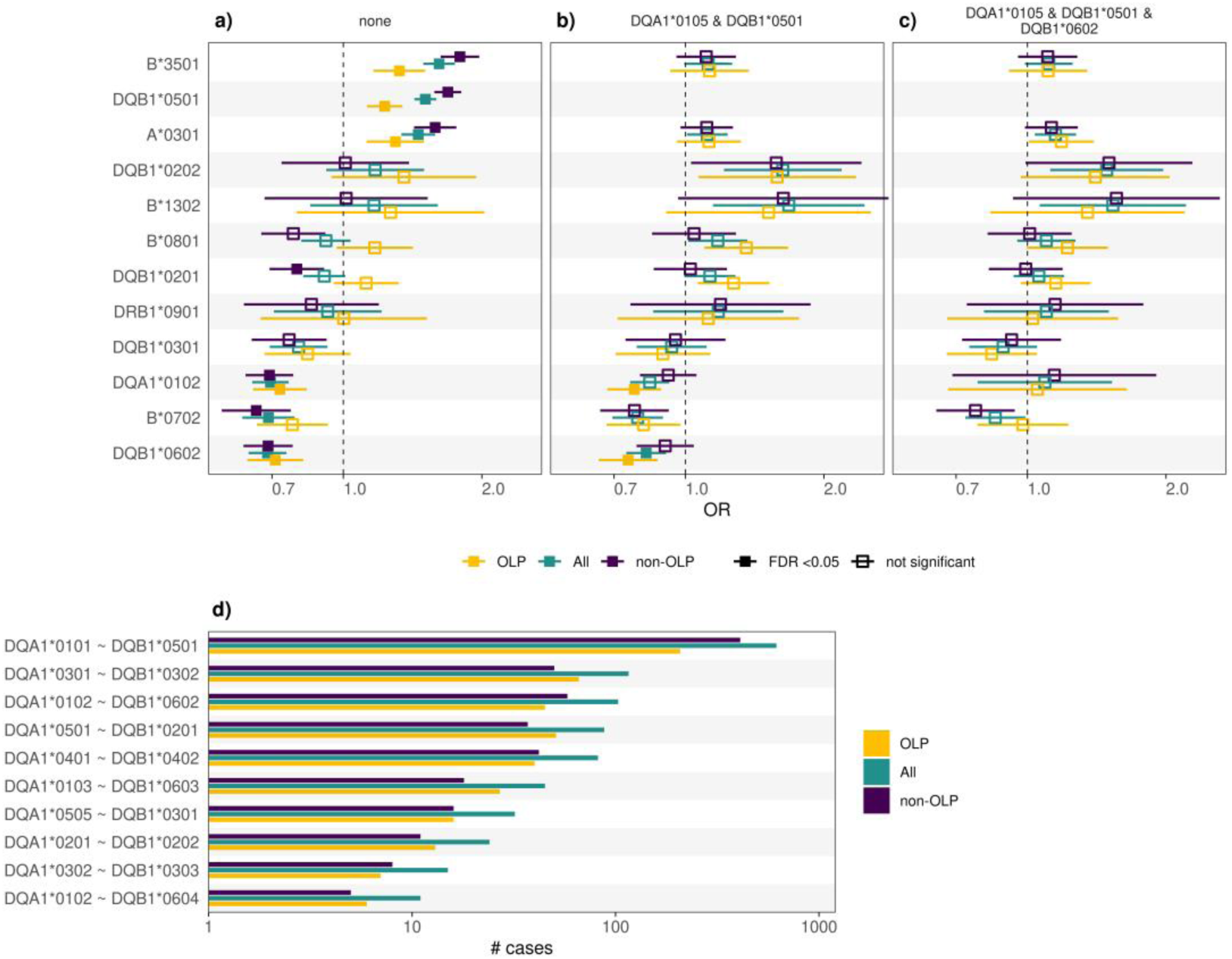
LP association analysis limited to DQA1 ∼ DQB1 homozygotic samples. **a)** Odds ratios (OR, x-axis) of top alleles from a model adjusted for basic covariates. **b)** As in (a), but with DQA1*01:05 and DQB1*05:01 allele dosages included as additional covariates. **c)** As in (a), but with DQA1*01:05, DQB1*05:01 and DQB1*06:02 allele dosages included as additional covariates. **d)** DQA1 ∼ DQB1 homozygous haplotypes (y-axis) and their corresponding numbers of cases available for analysis. Error bars in (a-c) represent 95% confidence intervals.

### *HLA-DQB1*05:01* effect increases susceptibility in combination with multiple *DQA1*01* alleles

Of note, the *DQB1*05:01* association was stronger than the *DRB1*01:01* association, suggesting a primarily DQ effect. Indeed, *DQB1*05:01* is found in multiple extended haplotypes, including the common *DRB1*01:01∼DQA1*01:01∼DQB1*05:01* haplotype but also other less common haplotypes *DRB1*01:02* or *DRB1*01:03*.

Based on known, strong HLA haplotypes in the Finnish population, a primary *DQA1*01:01/DQB1*05:01* heterodimer association explains that *DQB1*05:01* was more strongly associated than *DRB1*01:01* (Table 1). Beyond *DQB1*05:01*, both *DRB1*10:01* (OR=2.24) and *DQA1*01:05* (OR=2.31) showed a slightly larger effect size than *DRB1*01:01* and *DQA1*01:01* (OR=1.68) but a lower p-value, reflecting the fact the *DRB1*10:01* haplotype is less common in the Finnish (and other European ancestry) population (Table 1). Further support for *DQ* as the primary locus of susceptibility is given by the sequence similarity of *DQA1*01:01∼DQB1*05:01* and *DQA1*01:05∼DQB1*05:01*, which only differs by two amino acid replacements, one of them in the signal peptide (-7, V to M), one another at position 2 (D to G)).

These findings establish both *DQA1*01:05/DQB1*05:01* and *DQA1*01:01/DQB1*05:01* as primary susceptibility heterodimers in this disease, with stronger associations observed in non-OLP than OLP patients, likely because non-OLP represents a more pathogenetically homogeneous group (Table 1).

### *DRB1*15:01∼ DQA1*01:02∼DQB1*06:02* protective effect

A strong protective effect was observed at *DRB1*15:01∼DQA1*01:02∼DQB1*06:02* (Table 1). As this combination is almost exclusively associated with *DRB1*15:01∼DQA1*01:02∼DQB1*06:02* in Finns (and other European ancestries), it is impossible to differentiate a DR or a DQ effect; additional studies in African, Middle Eastern or South Chinese populations would be needed to differentiate DR and DQ effects, as LD of *DQA1*01:02∼DQB1*06:02* with *DRB1*15:01* is not absolute in these ethnic groups(11, 25). As will be shown later, however, the *DRB1*15:01∼ DQA1*01:02∼DQB1*06:02* protective effect is only present in *DQB1*05:01* negative patients (in *DQB1*05:01* positive patients, *DQA1*01:02∼DQB1*06:02* increases risk, see below), albeit the risk differs between *DQA1*01:01* vs *DQA1*01:05* (Figure 2).

**Figure 2.**
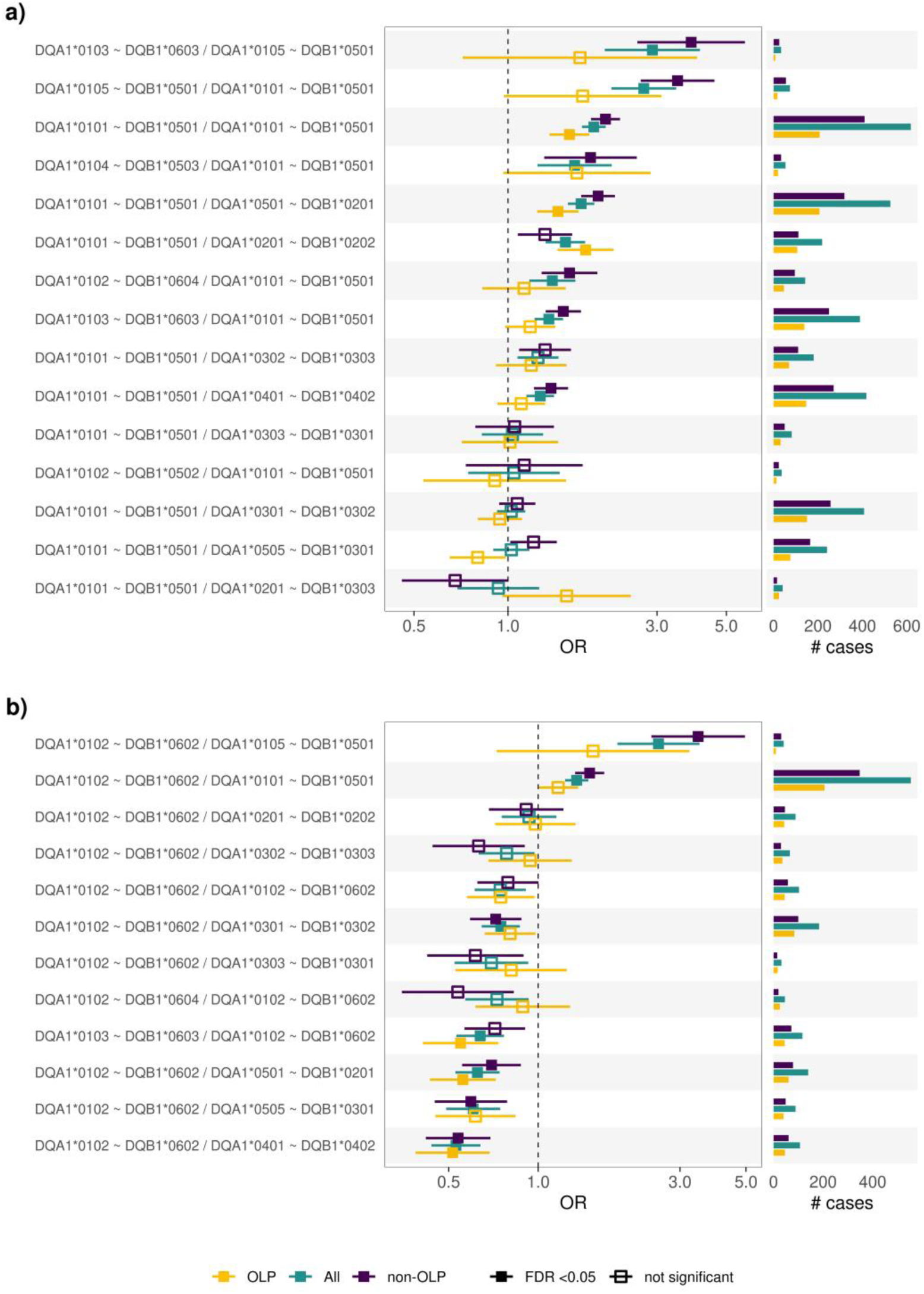
Selected DQA1 ∼ DQB1 diploid haplotypes and their LP association odds ratios (OR, x-axis), adjusted for Class I (A*02:01, A*03:01, B*08:01, B*13:02, B*07:02). **a)** Haplotypes containing the DQB1*05:01 allele. **b)** Haplotypes containing the DQB1*06:02 allele. Error bars represent 95% confidence intervals.

Other HLA class II effects were noted including an association with *DPB1*04:02* (increased susceptibility) and *DPB1*05:01* (protection) as well as weaker protective effects of *DRB1*13:01∼DQA1*01:03∼DQB1*06:03* and *DRB1*11:01∼DQA1*05:05∼DQB1*03:01*. These effects where no longer detected after controlling for the main effects as discussed later.

### Homozygous-only analysis of *DQA1∼DQB1* haplotypes confirms *DQB1*05:01* as the main risk factor and *DQB1*06:02* independently protective

A limitation in the above analysis is the potential role of *DQ* trans-heterozygote effects, which have been identified in other strongly *HLA-DQ* associated diseases such as celiac disease and narcolepsy(9–12). In some cases, novel heterodimers formed between α and β chains encoded on different chromosomes may increase disease susceptibility(12). In other cases, these trans-encoded heterodimers may be functionally inactive and reduce susceptibility by competing for heterodimerization with the primary susceptibility heterodimer encoded in cis(10, 11). To test this, we first conducted an *HLA-DQ* homozygous analysis only (Figure 1), an approach where *DQ* homozygous individuals are compared with each other while controlling for population stratification using principal component analysis (PCA). Here, the most significant *DQ* heterodimers are removed sequentially until nothing significant is left, an analysis called a relative predisposition effect analysis(26).

One important disadvantage is that the homozygote only analysis greatly reduces power, thus can only be conducted when large sample sizes are available, and for frequent haplotypes. For example, *DQA1*01:05∼DQB1*05:01* homozygotes are only expected at a frequency of 7 for 100,000 controls in this sample and perhaps ∼5 times more in patients, thus homozygosity effects cannot be observed even in our large sample.

This analysis (Figure 1, Supplementary Table 5) confirmed increased disease susceptibility in individuals homozygous for *DQA1*01:01∼DQB1*05:01*. Additionally, after conditioning on *DQB1*05:01* (Figure 1b), we observed an independent protective effect in *DQA1*01:02∼DQB1*06:02* homozygotes that persisted even after controlling for the primary susceptibility factors, indicating this protection operates independently of *DQA1*01∼DQB1*05:01*. Notably, this protective effect reached statistical significance only in the OLP subgroup and in the combined analysis of all cases, but not in the non-OLP subgroup where the effect was weaker.

### Further analysis on associated haplotypes as heterozygous shows trans-heterodimer effects

Having identified *HLA-DQA1*01:05/DQB1*05:01* and *DQA1*01:01/DQB1*05:01* heterodimers as the main susceptibility factors, and *DQA1*01:02∼DQB1*0602* as an independent protective haplotype, we next studied LP predisposition in heterozygous individuals with the DQ1 haplotypes (Figure 2).

We examined only combinations involving frequent haplotypes in the population and those associated with LP. Since the predisposing effect of *DQB1*05:01* and the protective effect of *DQB1*06:02* were found to be independent of each other, we analyzed the effects of *DQA1*01:01/5∼DQB1*05:01* and *DQA1*01:02∼DQB1*06:02* combinations separately. In all subsequent figures, we present data from OLP, non-OLP, and combined LP samples, noting that non-OLP generally shows stronger HLA associations. To eliminate potential confounding effects from class I alleles, these analyses were conducted both with and without controlling for *HLA-A*03:01*, *B*13:02*, and *B*08:01*—independent HLA Class I effects that we identified in subsequent analysis (detailed below).

As can be seen in Figure 2, a striking pattern of susceptibility was evident when various DQ1 heterozygous are present in trans of *DQB1*05:01* (panel a). For example, the *DQA1*01:03∼DQB1*06:03/DQA1*01:05∼DQB1*05:01* or *DQA1*01:02∼DQB1*06:02/DQA1*01:05∼DQB1*05:01* heterozygote combinations carried as much susceptibility as *DQA1*01:01∼DQB1*05:01/ DQA1*01:05∼DQB1*05:01* (containing two high susceptibility haplotypes). This indicates that trans *DQA1*01:02* or *03/DQB1*05:01* heterodimers carry as much susceptibility as *DQA1*01:01/DQB1*05:01* in predisposing to disease (*DQA1*01:03∼DQB1*06:03* or *DQA1*01:02∼DQB1*06:04* homozygotes have no effects or insignificant weak protective effects in the homozygous only analysis). This is also confirmed by the fact *DQA1*01:03∼DQB1*06:03/DQA1*01:01∼DQB1*05:01* (panel a) and *DQA1*01:02∼DQB1*06:02/DQA1*01:01∼DQB1*05:01* (panel b) carries similar or slightly lower risk as *DQA1*01:01∼DQB1*05:01/DQA1*01:01∼DQB1*05:01* homozygotes. Similarly, the genotype *DQA1*01:04∼DQB1*05:03, DQA1*01:01∼DQB1*05:01* has a similar risk as *DQA1*01:01∼DQB1*05:01* homozygotes.

Overall, results suggest that *DQA1*01:02, DQA1*01:03/DQB1*05:01* heterodimers carry as high a susceptibility risk as *DQA1*01:01/DQB1*05:01*, but significantly lower risk than *DQA1*01:05/DQB1*05:01*. Of note, *DQB1* is also likely to have some influence, as *DQA1*01:02∼DQB1*06:04* and *DQA1*01:02∼DQB1*05:02* in trans of *DQA1*01:01∼DQB1*05:01* reduces somewhat susceptibility. Further, *DQA1*01:01∼DQB1*05:01* with *DQA1*01:04∼DQB1*05:03* also has a somewhat lower risk, and as *DQA1*01:04* and *DQA1*01:05* are molecularly equivalent, being only distinct in exon 3 at position 199 where *DQA1*01:04* carries a T instead of an A in all other *DQA1* subtypes (Supplementary Figure 1)

Independent of this, *DRB1*15:01∼DQA1*01:02∼DQB1*06:02* was found to protective in all other heterozygous combinations, but this protective effect was only found in *DQB1*05:01* negative patients (panel b), suggesting heterogeneity of effects in *DQB1*05:01* positive vs negative patients.

### Conditional HLA allele associations reveal non-OLP vs OLP differences

Following the findings described above, analyses were repeated after controlling for the main HLA class II effects, *DQA1*01:05*, *DQB1*05:01* (and *DQA1*01:01*), followed by *DQB1*06:02*, revealing a disappearance of the *B*35:01* and *C*04:01* effects, indicating that these HLA class I effects were secondary to LD with *HLA-DR* and *DQ*. All these findings were made overall and in both non-OLP and OLP subtypes, with stronger effects in non-OLP (Figure 3).

**Figure 3.**
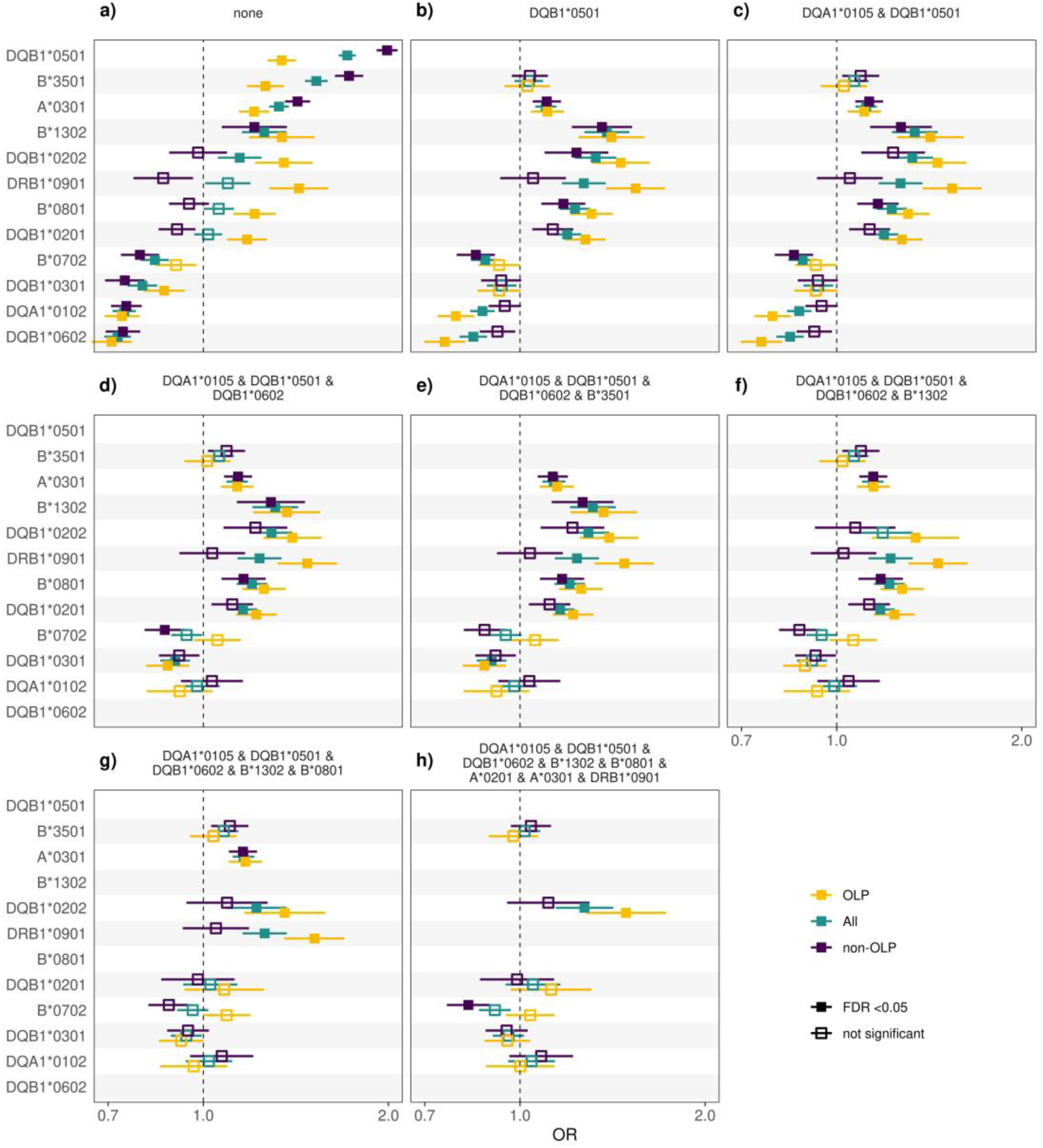
Results of LP association analysis for top HLA alleles. **a-h)** Odds ratios (OR, x-axis) of a conditioning chain starting from basic covariates and progressively adding HLA alleles as additional covariates to the logistic regression model. The alleles included as additional covariates are given on top of each panel. Error bars represent 95% confidence intervals.

As shown n Figure 3, it can be noted that controlling for *DQA1*01:05*, *DQB1*05:01*, and *DQB1*06:02* eliminates most HLA class II effects, except for weak predisposing effects of *DRB1*09:01* and *DQB1*02:02* that was more preeminent in OLP, the subtype least associated with *DQB1*05:01* and *DQB1*06:02*. *DRB1*09:01*, a frequent Asian subtype, is present in the context of the *DRB1*09:01∼DQA1*03:02∼DQB1*03:03* haplotype; further analysis would be needed to establish whether the effect is *DR* or *DQ* mediated.

The *DQB1*02:02* effect, predominant in OLP cases, was likely mediated by *DQA1*02:01/DQB1*02:02* heterodimers rather than by *DRB1*07:01*. Indeed, *DRB1*07:01* is commonly associated with *DQA1*02:01∼DQB1*02:02* and *DQA1*02:01∼DQB1*03:03* in the general population. As *DQB1*02:02* is more strongly associated than *DRB1*07:01* or *DQA1*02:01* (OR 1.35 vs. 1.24; Supplementary Data, basic allele analysis), this effect is more likely mediated by *DQA1*02:01/DQB1*02:02* than by *DRB1*. Further, *DQB1*02:01*, a subtype differing only in exon 3 and functionally equivalent to *DQB1*02:02*, present in the *DRB1*03:01∼DQA1*05:01∼DQB1*02:01* was not associated. Studies in African populations in which a haplotype bearing *DRB1*09:01-DQB1*02:02* is common would be particularly informative to confirm these possible additional susceptibility factors.

These results confirm that LP is primarily associated with *DQA1*01:05/DQB1*05:01*, *DQA1*01:01/DQB1*05:01* (strongest in non-OLP), and very weakly with *DQA1*02:01/DQB1*02:02* (stronger in OLP) and *DRB1*09:01∼DQA1*03:02∼DQB1*03:03* (only in OLP). It also displays a strong protective association with *DRB1*15:01∼DQA1*01:02∼DQB1*06:02* (strongest in non-OLP).

Following controlling for these HLA-class II effects, a predisposing effect at *A*03:01* (both OLP and non OLP), *B*13:02* (stronger in non-OLP) and *B*08:01* (stronger in OLP) remained (Figure 3). In contrast, effects at *B*35:01* and *C*04:01* found in the uncorrected sample association (Table 1) disappeared, all explained by LD (Supplementary Figure 3).

### Conditioning MHC SNP effects on *DQA1*01:05, DQA1*01:01, DQB1*05:01* and *DQB*06:02* shows that the association signal is explained by HLA alleles

We next explored how the GWAS SNP signals observed in the HLA region changed following conditioning on *DQA1*01:05, DQB1*05:01* and *DQB*06:02*, which represent most of the HLA class II signal in LP. As can be seen in Figure 4, although a weak signal may remain in the HLA class II region, additional signals were found in HLA Class I as found in other *DQ* associated diseases such as Type 1 diabetes(27, 28), celiac disease(29) or narcolepsy(9). Unlike in these other diseases, no effect is observed in the *HLA-DP* region(9, 27, 28, 30). The HLA class I signal disappeared when controlling for *A*03:01, HLA-B*13:02* and *B*08:01*, indicating that our analysis considered most effects present within the HLA region.

**Figure 4.**
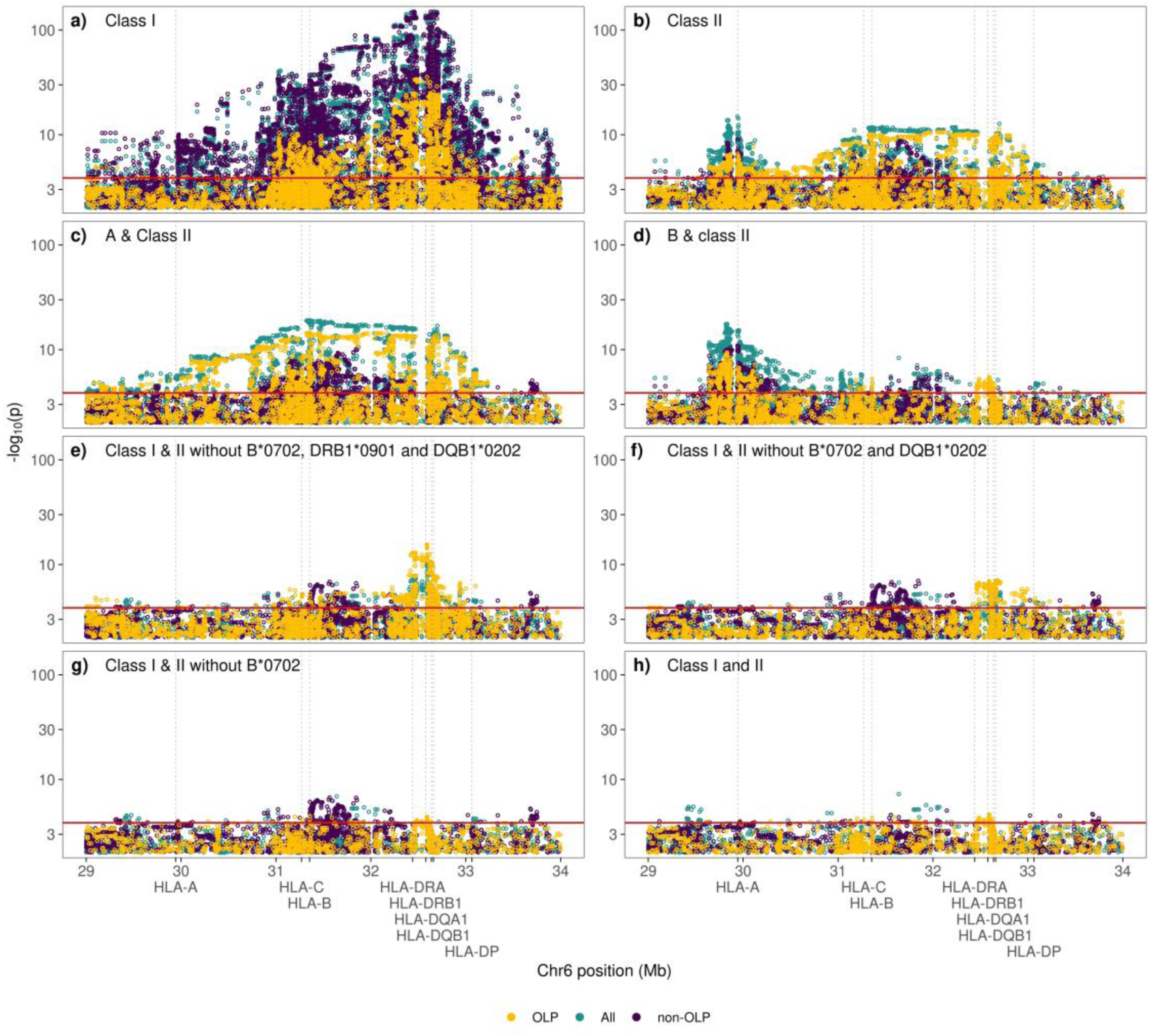
Manhattan plots of LP association p-values of MHC region SNPs on chromosome 6. **a-h)** Analyses adjusted for basic covariates and HLA alleles as indicated on top of each panel. Class I: *A*02:01*, *A*03:01*, *B*08:01*, *B*13:02*, *B*07:02*; Class II: *DQA1*01:05*, *DQB1*02:02*, *DQB1*03:01*, *DQB1*05:01*, *DQB1*06:02*, *DRB1*09:01*; A: *A*02:01*, *A*03:01*; B: *B*08:01*, *B*13:02*, *B*35:01*, *B*07:02*. The red horizontal line indicates the genome-wide significance threshold (p < 5 × 10^−8^)

## Discussion

This study furthers our understanding of HLA effects in LP and illustrates the complexity of *HLA-DQ* associated diseases. As previously reported, we found that the strongest HLA effects in LP are a *DQB1*05:01* predisposing effect and an independent *DQB1*06:02* protective effect, and that both effects are stronger in non-OLP versus OLP. This suggests that non-OLP (typically cutaneous), a disease affecting 0.2% to 1% worldwide, is autoimmune-like and has a more homogeneous etiology(1, 2). Nonetheless, it is worth noting that even for non-OLP cases, *DQB1*05:01* frequency was only 31% vs 19% in controls (Table 2), suggesting complexity and heterogeneity. Further, only in *DQB1*05:01* negative patients was *DQB1*06:02* protective, thus at least three separate subtypes of disease are most likely present: *DQB1*05:01* positive, both *DQB1*05:01* and *DQB1*06:02* negative and *DQB1*05:01* negative but *DQB1*06:02* positive. A clinical comparison of these subtypes could reveal clinical and prognostic differences of interest to the dermatologic communities.

Oral lichen planus (prevalence 1%)(31), in contrast, although sharing the same pathophysiology in a significant number of cases (approximately a third considering differences in odds ratios), is more heterogeneous. Considering the overlapping HLA association in OLP and non-OLP, it is likely that OLP includes at least 70% of cases with a pathophysiology that does not involve the same autoimmune process as non-OLP. Some of these cases have a weak HLA association with *DQA1*02:01/DQB1*02:02* and *DRB1*09:01*. *DRB1*09:01* predominates in Asia and thus could be best replicated in non-European ancestries. The association with *DRB1*09:01∼DQA1*03:02∼DQB1*03:03* is also present in rare *DQB1*06:02* positive OLP cases (Figure 2), suggesting a real subtype more frequently oral but also possibly cutaneous. In contrast, the *DQB1*02:02* associated allele was not increased in *DQB1*06:02* positive non-OLP cases (Figure 2), suggesting those cases to be more non-oral.

The weakly HLA-associated OLP cases may either share a distinct autoimmune basis or have infectious or other causes. Indeed, many diseases of diverse pathophysiology, including infections(32, 33), cancer(34, 35) and even neurodegenerative diseases(36) have weak HLA associations. Here too, additional clinical correlation and studying potential triggers stratified by HLA subtypes may be informative. Lichen planus (more cutaneous than oral), has been suggested to be associated with and triggered by hepatitis C(5–7), a virus whose clearance is modulated by *HLA-DR* and *DQ*, notably *DRB1^∗^01:01*, *DQA1^∗^03:03/DQB1^∗^03:01*, *DQA1^∗^05:05/DQB1^∗^03:01* for clearance and *DQA1^∗^01:02/DQB1^∗^06:02* and *DQA1^∗^05:01/HLA-DQB1^∗^02:01* for persistence(37). Small studies have also suggested differences in HLA association in some LP subgroups, such as *DQB1*02:02* in the vulvovaginal gingival form(38). Of additional interest, OLP carries a higher documented cancer risk(39), which would be interesting to explore in cases with *DQB1*05:01, DQB1*06:02*, *DQB1*02:02* and *DRB1*09:01*.

More detailed analysis offered additional information on the molecular nature of the *DQB1*05:01* association. Because non-OLP is more strongly HLA-associated, the signal was easiest to dissect in this subgroup. Strikingly, *DQB1*05:01*, the dominant effect, not only exhibited an approximate doubling of predisposing effects for *DQA1*01:01∼DQB1*05:01* homozygous vs heterozygous, as reported in other HLA associated diseases(40–44), but also showed a unique pattern of increased heterozygosity with any other *DQA1*01* alleles in trans.

Two effects were noted. First, as expected, increased susceptibility of *DQA1*01:05∼DQB1*05:01* (*DRB1*10*) was noted in trans of any haplotype versus the *DQA1*01:01∼DQB1*05:01* (*DRB1*01*) version. As an example, *DQA1*01:01∼DQB1*05:01*/ *DQA1*01:05∼DQB1*05:01* carried higher susceptibility than *DQA1*01:01∼DQB1*05:01* homozygotes in non-OLP (*DQA1*01:05∼DQB1*05:01* homozygotes are too rare to be tested) (Figure 2).

It can be speculated that *DQA1*01:05* vs. *DQA1*01:01* could contribute distinct physiological effects to explain the observed higher risk for *DQA1*01:05/DQB1*05:01* in non-OLP, for example better binding of a common epitope in the presence of *DQA1*01:05*. A similar finding exists in another *DQB1*05:01* associated disease, anti inglon-5, where *DQA1*01:05∼DQB1*05:01* is much more strongly associated than *DQA1*01:01∼DQB1*05:01*, and where the risks had the following order: *DQA1*01:05∼DQB1*05:01* > *DQA1*01:01∼DQB1*05:01* > *DQA1*01:04∼DQB1*05:03*(24). As demonstrated by our results, analysis of homozygous and heterozygous DQ1 carriers revealed that any *DQA1*01* allele (*DQA1*01:01, DQA1*01:02, DQA1*01:03, DQA1*01:04*, or *DQA1*01:05*) in combination with *DQB1*05:01* increases disease susceptibility. This suggests all *DQA1*01/DQB1*05:01* heterodimers predispose individuals to LP, although with varying degrees of effect—notably, *DQA1*01:05/DQB1*05:01* demonstrating a stronger association than *DQA1*01:01/DQB1*05:01*.

Second and more surprisingly, the presence of *DQA1*01:03* and *DQA1*01:02* in trans of *DQA1*01:05∼DQB1*05:01* increased susceptibility almost as much as *DQA1*01:01∼DQB1*05:01* homozygous (Figure 2). Intriguingly, in trans with *DQB1*01:05∼DQB1*05:01*, the effect of *DQA1*01:03* and *DQA1*01:04* was equivalent or stronger than *DQA1*01:01∼DQB1*05:01* homozygous, while *DQA1*01:03* and *DQA1*01:04* in the presence of *DQA1*01:01∼DQB1*05:01*, had slightly lower predisposition (Figure 2). These findings demonstrate in LP, all *DQA1*01/DQB1*05:01* combinations independent of *DQA1*01* diversity are predisposing. *DQA1*01:05/DQB1*05:01* has the highest effect, followed by *DQA1*01:01/DQB1*05:01* and then *DQA1*01:03/DQB1*05:01*, *DQA1*01:04/DQB1*05:01*, and *DQA1*01:02/DQB1*05:01*.

Of note, the exact ranking of predisposition for these DQ5 heterodimers was difficult to determine without a full understanding of the epitope(s) involved and its/their binding properties to *HLA-DQ*. Indeed, it is likely that the “other” *DQB1*05* or *DQB1*06* alleles of these haplotypes are not entirely neutral, and as these can also pair with *DQA1*01* alleles, *DQB1* allele competition could occur. This could reduce *DQA1*01/DQB1*05:01* abundance and susceptibility, a phenomenon observed in narcolepsy, although primarily with *DQA1*01* alleles(9–11). In favor of this, *DQA1*01:04* is likely molecularly equivalent to *DQA1*01:05*, as it differs in exon 3 at position 199 (A->T), and area of *DQA1* that is not known to change peptide binding. Despite this however, *DQA1*01:04∼DQB1*05:03* does not rank high in predisposition in trans to *DQA1*01:01∼DQB1*05:01*, suggesting that *DQB1*05:03* could reduce the effect of *DQA1*01:04∼DQB1*05:01*, a competing effect.

Nonetheless, although *DQB1*05:01* differs from *DQB1*06* at multiple positions, it only differs at position β57 from *DQB1*05:02* (V->S) and *DQB1*05:03* (V->D), two alleles that are not associated with LP (Supplementary Figure 1). The β57 position is critical to peptide binding at P9, the most C terminal portion of the presented peptide. Similarly, *DQA1*01* alleles differ from each other at positions α2, α25, α34 and α41, positions that mostly interact with P1 and less so with P4(45). This suggests that an important peptide presented in LP must involve conserved binding through P6 and P9, while being more tolerant at N terminal positions P1 and P4. This prediction will only be verifiable once the autoantigen is identified.

The independent protective effects of *DRB1*15:01∼DQA1*01:02∼DQB1*06:02*, which is only visible in *DQB1*05:01* negative individuals, is more difficult to explain; protective effects such as that of this haplotype in Type 1 diabetes are still largely unexplained(46). It may suggest the existence of a separate subtype of the disease or be also present in *DQB1*05:01* positive effect but superseded by the stronger risk effect of *DQA1*01:02/DQB1*05:01* on the disease. Studying this and similar effects in subjects with African ancestry, where *DRB1*12:01∼ DQA1*01:05∼DQB1*0501* and *DRB1*11:01∼DQA1*01:02∼DQB1*06:02* haplotypes are common, could help map this effect to DQ versus DR, although large numbers will be required as the protective association in lichen planus is not extremely high.

Besides HLA Class II effects, clear independent HLA class I effects were also noted with *A*03:01*, *B*13:02* and *B*08:01*, present in both non-OLP and OLP, as well as protective effects of *B*07:02* in OLP only. Associations of HLA class I alleles independent of class II allele associations is common to most autoimmune diseases such as Type 1 diabetes(47), multiple sclerosis(48), and narcolepsy(9). The role of HLA class I in LP is in line with the well-known observation that lesions in this disease involve both CD4 and CD8 T cells(4, 49). Most likely, although the antigen(s) is (are) unknown in this disease, CD4+ T cell help and involvement of CD8+ T cells is crucial. This is also revealed by the involvement of other immune loci, although interestingly these point to a role of eosinophils(8), cells that can also be HLA-Class II positive.

Unlike other autoimmune diseases however, autoantibodies have not been described, although to our knowledge, no systematic search has been initiated.

The pattern of HLA associations identified in this study provides valuable clues for epitope discovery in LP. The observation that all *DQA1*01/DQB1*05:01* heterodimers confer susceptibility—with varying degrees of risk—suggests that specific structural features of these molecules are critical for presenting pathogenic peptides. Particularly informative is our finding that *DQB1*05:01* confers risk while the closely related *DQB1*05:02* and *DQB1*05:03* alleles do not, differing only at position β57 (V→S/D), which influences peptide binding at P9. Similarly, the variable risk among different *DQA1*01* subtypes (with *DQA1*01:05* conferring the highest risk) points to the involvement of polymorphic residues at positions α2, α25, α34, and α41, which primarily affect binding at P1 and P4. These findings suggest that putative LP-associated epitopes likely maintain conserved anchoring at P6 and P9 positions while exhibiting greater tolerance for variation at N-terminal positions. This molecular fingerprint could significantly narrow the search for viral or autoantigens in LP, enabling targeted approaches that focus on peptides with these specific binding characteristics. Future studies employing in silico peptide binding predictions, followed by experimental validation with T cell assays from patients stratified by HLA risk genotypes, may successfully identify the elusive triggering antigen(s) in this complex disorder.

In summary, building upon the study by Reeve et al.(8), we have fine-mapped and characterized HLA association effects in LP. As observed in many other diseases primarily associated with HLA class II alleles, we identified complex effects within the HLA class II region along with additional, weaker effects in HLA Class I. Most of these associations can be attributed to amino acid polymorphisms that alter peptide binding rather than regulatory polymorphisms. A particularly notable feature of this disease is that the predisposing effect of *DQB1*05:01* remains largely invariant regardless of *DQA1*01* subtype. In this respect, LP joins other *HLA-DQ* associated diseases (narcolepsy, celiac disease, and anti-IgLON5 encephalitis) where strong trans DQα/β heterodimerization effects have been demonstrated to play a crucial role, necessitating specialized statistical modeling approaches beyond conventional genetic analysis.

## Data Availability

FinnGen summary statistics data are available at https://r12.finngen.fi/. All data produced in the present study are available upon reasonable request to the authors.

## Data and code availability

Analysis scripts are available in GitHub at https://github.com/FRCBS/LP_HLA. HLA allele association analysis summary statistics are available as Supplementary Data file. MHC region SNP association analysis summary statistics are available at https://drive.google.com/drive/folders/1k1ygQIfrXgD8CkO-djw4ykaFB_NgwRAV. FinnGen summary statistics data are available at https://r12.finngen.fi/.

## Acknowledgements

We want to acknowledge the participants and investigators of the FinnGen study. The FinnGen project is funded by two grants from Business Finland (HUS 4685/31/2016 and UH 4386/31/2016) and the following industry partners: AbbVie Inc., AstraZeneca UK Ltd, Biogen MA Inc., Bristol Myers Squibb Inc. (and Celgene Corporation & Celgene International II Sàrl), Genentech Inc., Merck Sharp & Dohme LCC, Pfizer Inc., GlaxoSmithKline Intellectual Property Development Ltd., Sanofi US Services Inc., Maze Therapeutics Inc., Johnson&Johnson Innovative Medicine Inc., Novartis AG, Boehringer Ingelheim International GmbH and Bayer AG. Following biobanks are acknowledged for delivering biobank samples to FinnGen: Auria Biobank (www.auria.fi/biopankki), THL Biobank (www.thl.fi/biobank), Helsinki Biobank (www.helsinginbiopankki.fi), Biobank Borealis of Northern Finland (https://www.ppshp.fi/Tutkimus-ja-opetus/Biopankki/Pages/Biobank-Borealis-briefly-in-English.aspx), Finnish Clinical Biobank Tampere (www.tays.fi/en-US/Research_and_development/Finnish_Clinical_Biobank_Tampere), Biobank of Eastern Finland (www.ita-suomenbiopankki.fi/en), Central Finland Biobank (www.ksshp.fi/fi-FI/Potilaalle/Biopankki), Finnish Red Cross Blood Service Biobank (www.veripalvelu.fi/verenluovutus/biopankkitoiminta), Terveystalo Biobank (www.terveystalo.com/fi/Yritystietoa/Terveystalo-Biopankki/Biopankki/) and Arctic Biobank (https://www.oulu.fi/en/university/faculties-and-units/faculty-medicine/northern-finland-birth-cohorts-and-arctic-biobank). All Finnish Biobanks are members of BBMRI.fi infrastructure (https://www.bbmri-eric.eu/national-nodes/finland/). Finnish Biobank Cooperative -FINBB (https://finbb.fi/) is the coordinator of BBMRI-ERIC operations in Finland. The Finnish biobank data can be accessed through the Fingenious® services (https://site.fingenious.fi/en/) managed by FINBB.

